# The impact of COVID-19 lockdown on a cohort of adults with recurrent major depressive disorder from Catalonia: a decentralized longitudinal study using remote measurement technology

**DOI:** 10.1101/2023.01.24.23284906

**Authors:** R. Lavalle, E. Condominas, JM Haro, I. Giné-Vázquez, R Bailon, E Laporta, E Garcia, S Kontaxis, G. Riquelme, F. Lombardini, A. Preti, MT Peñarrubia-María, M. Coromina, B. Arranz, E. Vilella, E. Rubio, RADAR-MDD-Spain, F. Matcham, F Lamers, M. Hotopf, BWJH Penninx, P. Annas, V Narayan, S. Simblett, S Siddi, the RADAR-CNS consortium

**Affiliations:** Dipartimento di neuroscienze, Università degli studi di Torino, Italia; Parc Sanitari Sant Joan de Déu, Fundació Sant Joan de Déu, CIBERSAM,Universitat de Barcelona, Barcelona, Spain; Aragón Institute of Engineering Research (I3A), University of Zaragoza, Zaragoza, Spain; Centros de investigación biomédica en red en el área de bioingeniería, biomateriales y nanomedicina (CIBER-BBN), Madrid, Spain; Microelectrónica y Sistemas Electrónicos, Universidad Autónoma de Barcelona, Spain; Health Technology Assessment in Primary Care and Mental Health (PRISMA) Research Group, Parc Sanitari Sant Joan de Deu, Institut de Recerca Sant Joan de Deu, St Boi de Llobregat, Catalunya, Spain; Unitat de Suport a la Recerca Regió Metropolitana Sud, Fundació Institut Universitari per a la recerca a l’Atenció Primària de Salut Jordi Gol i Gurina (IDIAPJGol), Barcelona, Catalonia, Spain; Hospital Universitari Institut Pere Mata, Reus, Spain; Institut d’Investigació Sanitària Pere Virgili-CERCA, Reus, Spain; Universitat Rovira i Virgili, Reus, Spain; Centro de investigación biomédica en red en salud mental, CIBERSAM-Instituto de Salud Carlos III, Madrid, Spain; King’s College London, Institute of Psychiatry, Psychology and Neuroscience, UK; School of Psychology, University of Sussex, East Sussex; Amsterdam UMC location Vrije Universiteit Amsterdam, Department of Psychiatry, Boelelaan 1117, Amsterdam, The Netherlands; Amsterdam Public Health, Mental Health program, Amsterdam, the Netherlands; H. Lundbeck A/S, Valby, Denmark; Research and Development Information Technology, Janssen Research & Development, LLC, Titusville, NJ, USA; https://radar-cns.org/

**Keywords:** SARS-CoV-2, Depression, Anxiety, Lockdown, Quarantine, Remote measurement technology, Decentralized study, Spain

## Abstract

**Background:** The present study analyzes the effects of each containment phase of the first COVID-19 wave on depression levels in a cohort of adults with a history of major depressive disorder (MDD).

**Methods:** This analysis is part of the Remote Assessment of Disease and Relapse-MDD (RADAR-MDD) study. Individuals included had a diagnosis of DSM-5 major depressive disorder (MDD), at least two episodes of major depression (MDE), one of them in the previous two years. Depression was evaluated with the Patient Health Questionnaire-8 (PHQ-8) and anxiety with the Generalized Anxiety Disorder-7 (GAD-7). A total of 121 participants recruited from Catalonia were registered from November 1, 2019, to October 16, 2020. Levels of depression were explored across the phases (pre-lockdown, lockdown, four post-lockdown phases) of the restrictions imposed by the Spanish/Catalan governments. Then, a mixed model was fitted to estimate how depression varied over the phases.

**Results:** A small but statistically significant rise in the depressive severity was found during the lockdown and phase 0 (early post-lockdown), as compared with the pre-lockdown phase in this sample with a history of MDD. Those with low pre-lockdown depression experienced an increase in depression levels during the “new normality”. We observed a significant decrease in the depression levels during the “new normality” in those with high pre-lockdown depression, compared to the pre-lockdown period.

**Conclusion:** These findings suggest that COVID-19 restrictions impacted on the depression of individuals diagnosed with MDD, depending on their pre-lockdown depression severity. Furthermore, these subjects worsened when the restrictions were harder, during the lockdown and the early post-lockdown.

## INTRODUCTION

In January 2020, the World Health Organization declared a novel severe acute respiratory coronavirus disease caused by SARS-CoV2, the main etiological factor of the disease, called COVID-19, Coronavirus Disease 2019 (Na Zhu et al., 2020). Soon governments from different countries imposed stringent restrictions to fight COVID-19 diffusion, such as lockdown or self-isolation (Ren et al., 2020). On March 11, 2020, the Government of Catalonia introduced social distancing to fight the spread of COVID-19 (RESOLUCIÓ SLT/704/2020, d’11 de Març, 2020). The Spanish Government then established strict lockdown measures the day after the declaration of the State of Alarm, on March 14, 2020 (Disposición 4911, BOE n.°130, 2020). During this first wave of the pandemic restrictions were lifted gradually through four phases of the post-lockdown implemented by the Spanish government and supplementary measures of the local Catalan government (Table S1). During phase 0 (Disposición 4791, BOE n.°123, 2020; Disposición 7351, BOE n.°107, 2021) non-essential businesses were opened by appointment, and citizens were allowed to do outdoor physical activities by time slot based on age. With phase 1 (Ajuntament de Barcelona, 2020a; Orden SND/399/2020, de 9 de Mayo, BOE n.°4911, 2020; Disposición 4911, BOE n.°130, 2020; la CIUTAT Diari digital de proximitat, 2020), meetings with a maximum of 10 people were allowed; only outdoor spaces of bars and restaurants opened, as well as some spaces of culture, museums, and gyms; transfers to a second residence were permitted. Later, during phases 2 and 3 (Ajuntament de Barcelona, 2020b; betevé, 2020; Jefatura de Estado, 2020; SER, 2020b, 2020a) time slots were abolished, and bars and restaurants’ openings were extended even to the indoor areas, with limited capacity; shopping centers opened, public transport restarted working at 100% and the percentage of capacity in cinemas, theaters, museums increased. Finally, these relaxations culminated in a period of “new normality”, as the government named it, from June 18 until October 16, 2020, when the COVID-19 second wave started (RESOLUCIÓ INT/1433/2020, de 18 de Juny, 2020).

The benefits in the control of the infection may have been at the expense of significant psychological impact (Rubin & Wessely, 2020). In particular individuals with mental health problems were described as running a higher risk of being further psychologically impacted by the pandemic, compared to the general population (Hampshire et al., 2022; Kunzler et al., 2021; Rodríguez-Fernández et al., 2021). A previous study, conducted on a Spanish sample, showed that during the first phases of the post-lockdown, people with mental illness suffered more from depressive symptoms than healthy controls (Solé et al., 2021). As regards patients with Major depressive disorder (MDD), some authors (Hamm et al., 2020) found no significant variations of depressive symptoms in an American sample between before and during the lockdown, while others (Quittkat et al., 2020) described an increase in a German population.

However, many of the reported studies lack data referring to the pre-pandemic levels of symptoms (García-Álvarez et al., 2020; González-Blanco et al., 2020; Martinelli et al., n.d.; Solé et al., 2021). Moreover, many were based on cross-sectional surveys (Asmundson et al., 2020; Billen et al., 2020; Hao et al., 2020; Korsnes et al., 2020; Liu et al., 2020; Quittkat et al., 2020; Solé et al., 2021; Zhu et al., 2021). Although there is a large body of literature on the psychological effects of COVID-19, most of it was conducted in the general population (Castaldelli-Maia et al., 2021; García-Álvarez et al., 2020; Jin et al., 2021; Kunzler et al., 2021; Luo et al., 2020; Phiri et al., 2021; Ren et al., 2020; Salari et al., 2020), or included patients with different types of mental disorders in the same sample (Asmundson et al., 2020; Billen et al., 2020; Franchini et al., 2020; Frank et al., 2020; Hao et al., 2020; Iasevoli et al., 2021; Korsnes et al., 2020; Liu et al., 2020; Muruganandam et al., 2020; Pan et al., 2021; Zhu et al., 2021), or used non-standardized instruments to assess the mental status (Franchini et al., 2020; Muruganandam et al., 2020), or did not take into account the different kinds of restrictions across countries during the first wave of COVID-19, (Leightley et al., 2021).

To adequately understand the impact of the pandemic on depression severity, we need longitudinal studies able to include people with a recent history of MDD. Two previous publications (Leightley et al., 2021; Siddi, et al., 2022) using data from the Remote Assessment of Disease and Relapse-Major Depressive Disorder (RADAR-MDD) study (Matcham et al., 2019), explored the depression severity pre-, during, and post-lockdown periods in people with a history of MDD from Spain, UK and the Netherlands, and observed that patients who displayed significant depressive symptoms shortly before the COVID-19 outbreak decreased in their severity between pre- and during the lockdown (Leightley et al., 2021). However, Leightley et al. (2021) and Siddi et al. (2022) only considered three periods: pre-lockdown, lockdown, and post-lockdown, but the social restrictions during the post-lockdown were in many places lifted gradually and in different stages, which implies significant differences across countries. The regional and local impact of the COVID-19 crisis has been highly heterogeneous with significant implications for crisis management and policy responses (*The Territorial Impact of COVID-19: Managing the Crisis across Levels of Government*, n.d.). For example, Spanish government adopted more restrictive measures than the Netherlands, calculated by the stringency index (highest index per country Spain, Netherlands and UK 85.2, 78.7 and 79.6, respectively, during the period from February until the first of October, 2020) measured by the COVID-19 government response tracker (*OxCGRT*, n.d.). There is some evidence that policy stringency consisted in physical distancing protocols impeding familiar and meaningful forms of social connection, and more stringent COVID-19 policies were associated with poorer mental health (Aknin et al., 2022).

The present study focuses on one site/country of the RADAR-MDD project to make more detailed analyses on how specific containment measures affected the mental health of individuals with a history of MDD. We aim to explore the impact of the COVID-19 control restrictions on mental health in individuals with a history of MDD in Catalonia (the Spanish sample of the RADAR-MDD study was recruited in this Autonomous Community), adopting a new ‘pandemic-centric’ perspective. RADAR-MDD is part of the RADAR-CNS (Remote Assessment of Disease and Relapse - Central Nervous System) consortium (https://www.radar-cns.org/), which developed the open-source mHealth platform RADAR-Base (Ranjan et al., 2019) to collect longitudinal data using remote measurement technology (RMT), using active apps installed in a smartphone providing data on depressed mood, self-esteem, speech, and cognition; and passively (without the interactions with the participants) on heart rate, physical activity, sleep, and sociability throughout wearable device and passive app installed in a smartphone. In this study, we assessed the fluctuations of depression during the different phases of the first wave of the COVID-19 pandemic in relation to the limitations imposed by the national Spanish government and supplementary measures of the local policy of the Catalan authorities, by considering the depression and anxiety severity before the pandemic.

## METHOD

### 1. Study design and participants

The present study is based on data gathered in the RADAR-MDD project, (Matcham, et al., 2019; Matcham et al., 2022) a multi-center cohort study (Netherlands, Spain, and the UK) (https://www.radar-cns.org/) including people with a history of MDD who were evaluated from November 2017 to March 2021. The study was co-developed with service users in our Patient Advisory Board (PAB), who were involved in the choice of measures, the timing and issues of engagement and have also been involved in developing the analysis plan and a representative is author of this paper and critically reviewed it.

In this manuscript, we focused our analyses on the 155 participants with a history of MDD according to the DSM-5 criteria evaluated by clinicians from Catalonia and analyzed the data during the first wave of COVID-19, between November 1, 2019, and October 16, 2020. Of those 155 participants, 22 were not recorded during the periods of interest (from 01/11/19 to 16/10/20) and 12 missed records during the pre-lockdown period; thus, the final sample included 121 participants.

Additionally, some participants had missing values that referred to certain phases (see the supplementary materials for the number of observations in each phase, Table S2). No differences in the PHQ-8 pre-lockdown values of these patients with missing records in certain phases were observed (p>0.05) as compared to those without any missing data.

#### Pandemic phases

The phases and restrictions had a longer duration or were applied later depending on the location (Barcelona, Tarragona or Garraf). The dates of the restrictions are shown in Figure 1 and for a full description, see the supplementary materials (Table S1). On this basis, we focused on the following phases:

**Figure1.**
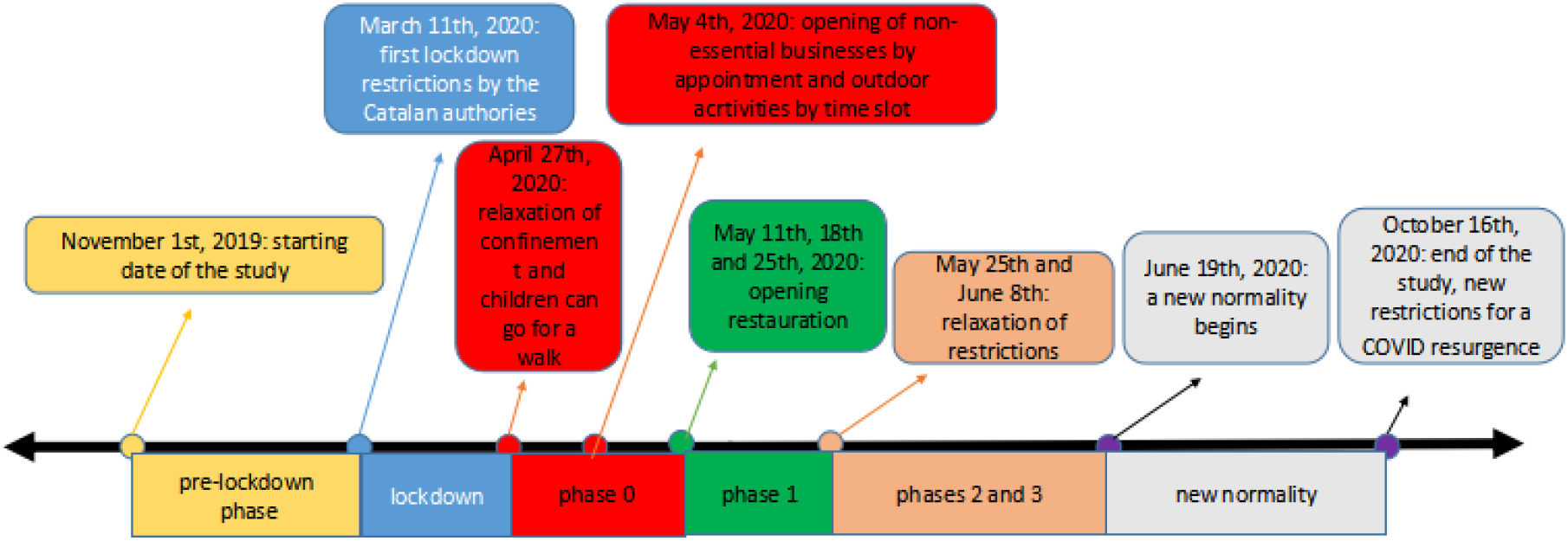
Description of the pre-, lockdown and post-lockdown phases.

- Pre-lockdown (from November 1, 2019, to March 10, 2020)
- Lockdown (from March 11, 2020, to April 26, 2020):

Later restrictions were lifted gradually through four phases of the post-lockdown:

- Phase 0 (from April 27, 2020, to May 10 or 17 or 24, 2020, depending on the location)
- Phase 1 (from May 11 or 18 or 25, 2020, to May 24 or June 7, 2020, depending on the location)
- Phases 2 and 3 (from May 25 or June 8, 2020, to June 18, 2020, depending on the location).
- “New” normality (from June 19, 2020, to October 16, 2020)

## 3. Measures

### 3.1 Depressive symptoms

Depression severity was assessed through the PHQ-8 (Patient Health Questionnaire-8) (Kroenke et al., 2009). Participants were required to answer this questionnaire every two weeks. The total score could range from 0 to 24, with increasing number meaning higher severity, a score of 10 was set as a cut-off (Kroenke et al., 2009), above which the severity of symptoms could be considered clinically relevant. Internal consistency was calculated on 3310 observations, with Cronbach’s alpha of 0.917.

### 3.2 Anxiety symptoms

Severity of anxiety was assessed using the Generalized Anxiety Disorder-7 item (GAD-7) scale (Spitzer et al., n.d.), which was collected at baseline and every 3 months. The GAD-7 score could vary from 0 to 21, with increasing number meaning higher severity, and a score of 10 was assumed as a cut-off (Spitzer et al., n.d.), above which the severity of symptoms could be considered clinically relevant. Internal consistency was calculated on 1178 observations, with Cronbach’s alpha of 0.971.

### 3.3 Sociodemographic variables

Gender, comorbidity with medical conditions, age, marital status, number of people living with, employment, income, and age of finishing education were also collected at baseline.

## 4. Statistical analysis

First, we described the sociodemographic characteristics of the sample and computed a descriptive analysis of the PHQ-8 values among the different lockdown phases. We then calculated each participant’s baseline depression and anxiety severity by taking the average of the PHQ-8 and GAD measures in the pre-lockdown period. Participants were grouped based on their pre-lockdown depression (PHQ-8≥ 10 or <10) and their pre-lockdown anxiety severity (GAD-7≥ 10 or <10). We analysed the variations of PHQ-8 in the different phases of the pandemic using the paired Wilcoxon text. In those participants with more than one PHQ-8 evaluation in the same phase, we computed the mean values and used them for the following analyses. We also applied a linear mixed model to analyse the course of depression levels over phases. To identify the covariate variables (sociodemographic variables) that would be included in the linear mixed model, we computed the forward stepwise method implemented with R based on the best Akaike information criterion (AIC) value. We also evaluated the effect size on pre-lockdown depression levels of each variable and added to the model those not selected by the stepwise procedure but with at least a medium effect (Cohen’s d ≥ 0.5 for categorical variables, and Spearman correlation coefficient ≥ 0.3 for continuous variables). To investigate how the pre-lockdown depression levels associated with variations in depression severity across phases, the model was stratified by pre-lockdown depression severity (PHQ-8 cutoff ≥ 10). Random effects of participants were incorporated as well and the model was adjusted using the maximum likelihood (ML) method, which provided correct estimation when data are not completely missing at random (Leightley et al., 2021). All statistical analyses were conducted using the nlme package in R (Pinheiro & Bates, D., DebRoy, S., Sarkar, D., 2021) and STATA 13.

## RESULTS

Most of the participants were females (66.9%), the median age was 58 years (IQR 52-64) (table 1). More than half of the individuals had high levels of anxiety (64.5%) and depression (PHQ-8) (67.8%) during the pre-lockdown phase.

**Table 1.** Characteristics of study population.

|  |  | (N=121) |
| --- | --- | --- |
| **Gender, N(%)** | Male | 40 (33,1%) |
|  | Female | 81 (66,9%) |
| **Comorbidity\*, N(%)** | Yes | 74 (61,2%) |
|  | No | 47 (38,8%) |
| **Age, Median (IQR)** |  | 58 (52-64) |
| **Marital status, N(%)** | with a partner | 68 (56,2%) |
|  | without a partner | 53 (43,8%) |
| **People living with N(%)** | alone | 22 (18,2%) |
|  | two | 40 (33,1%) |
|  | three | 37 (30,6%) |
|  | four or more | 22 (18,2%) |
| **Employment, N(%)** | employed | 43 (35,5%) |
|  | unemployed | 78 (64,5%) |
| **Income, N(%)** | < 15,000€ | 33 (27,3%) |
|  | 15,000€ - 24,000€ | 48 (39,7%) |
|  | > 24,000€ | 40 (33,1%) |
| **Age of finishing education, mean (SD)** |  | 17,6 (4,95) |
| **Pre-lockdown PHQ-8** | Mean | 12.90 |
|  | Median (IQR) | 13 (10.2) |
| **Pre-lockdown GAD-7** | Mean | 10.76 |
|  | Median (IQR) | 10 (5.2) |
| **Pre-lockdown PHQ-8, N(%)** | PHQ-8 <10 | 39 (32.2%) |
|  | PHQ-8 ≥ 10 | 82 (67.8%) |
| **Pre-lockdown GAD-7, N(%)** | GAD-7 < 10 | 43 (35.5%) |
|  | GAD-7 ≥ 10 | 78 (64.5%) |

**Note:** number of participants and proportion in parenthesis are displayed for categorical variables. Median and IQR or mean and standard deviation (SD) are shown for continuous variables. The values of income correspond to the gross salary per year. Unemployment is also considered retirement or work leave
\* Comorbidity the simultaneous presence of other diseases or medical conditions in a patient apart of the depression.

Figure 2 shows descriptive analysis of the depression severity measured by PHQ-8 values in the different phases of the pandemic.

**Figure 2.**
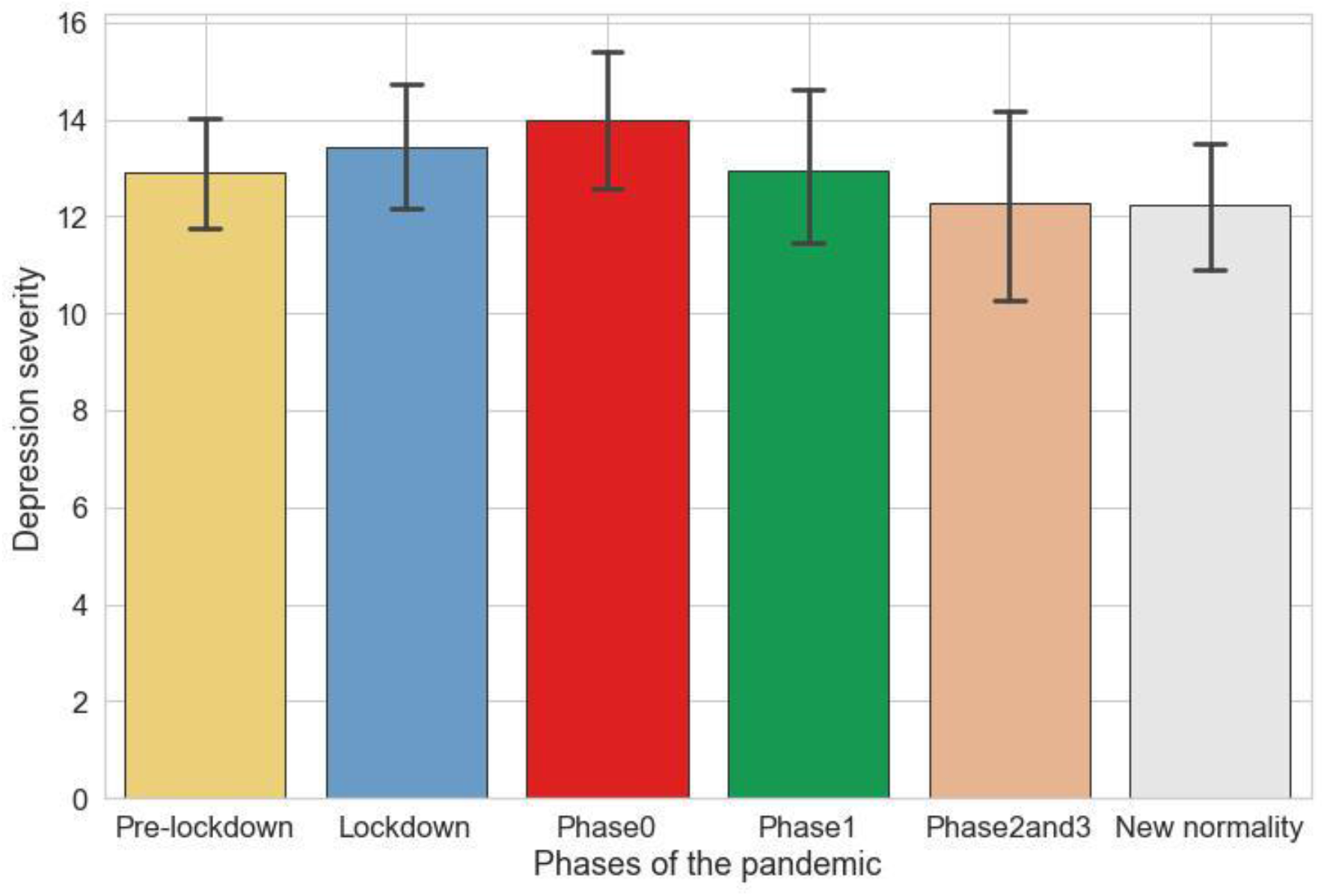
Bar plot of PHQ8 score during the pandemic phases. Each bar corresponds to the mean and confidence intervals of depression severity assessed by PHQ-8 during each phase of the pandemic.

Using the paired Wilcoxon test, a statistically significant increase in the depression severity (PHQ-8 score) was observed in phase 0 compared to the pre-lockdown (p≤0.05) and new-normality (p≤0.01) phases, while a significant decrease was found in new-normality as compared with lockdown (*p*≤0.05).

A linear mixed model (Table 2) was fitted to explore the association between phases and anxiety and depression severity, adjusted for the covariates (gender, age, income and number of people living with, and comorbidity with other medical conditions). A significant rise in depressive symptoms was found during the lockdown (β = 0.866, CI 95% [0.430-1.303], *p*≤0.001) and phase 0 (β = 1.135, CI 95% [0.560-1.711], *p*≤0.001), as compared with pre-lockdown phase. Depression severity was higher in patients with high pre-lockdown anxiety (GAD-7≥ 10) (β = 7.955, CI 95% [6.240-9.670], *p*≤0.001), than in those with low pre-lockdown anxiety symptoms (GAD-7<10).

**Table2.** Depression severity across phases compared to pre-lockdown levels measured with the PHQ-8 (Linear mixed model)

|  |  | Coef. | CI (95%) | p-value |
| --- | --- | --- | --- | --- |
| Phase | pre-lockdown | Ref | - | - |
|  | lockdown | 0.866 | [0.430 to 1.303] | ≤0.001 |
|  | phase0 | 1.135 | [0.560 to 1.711] | ≤0.001 |
|  | phase1 | -0.004 | [-0.709 to 0.700] | 0.990 |
|  | phase2-3 | 0.098 | [-0.770 to 0.967] | 0.824 |
|  | new normality | -0.099 | [-0.437 to 0.238] | 0.562 |

**Note: Mixed model coefficient without interaction and** adjusted for the covariates: gender, age, income and number of people living with, and comorbidity with other medical conditions.

We then stratified participants into two groups’: one contained subjects with high pre-lockdown depression (82 participants and 1048 observations), the other subjects with low pre-lockdown depression (39 participants and 547 observations). In this way, we aimed to verify whether the association between the phases and depression severity depend on the pre-lockdown depression severity (see Figure 3 and 4).

**Figure 3.**
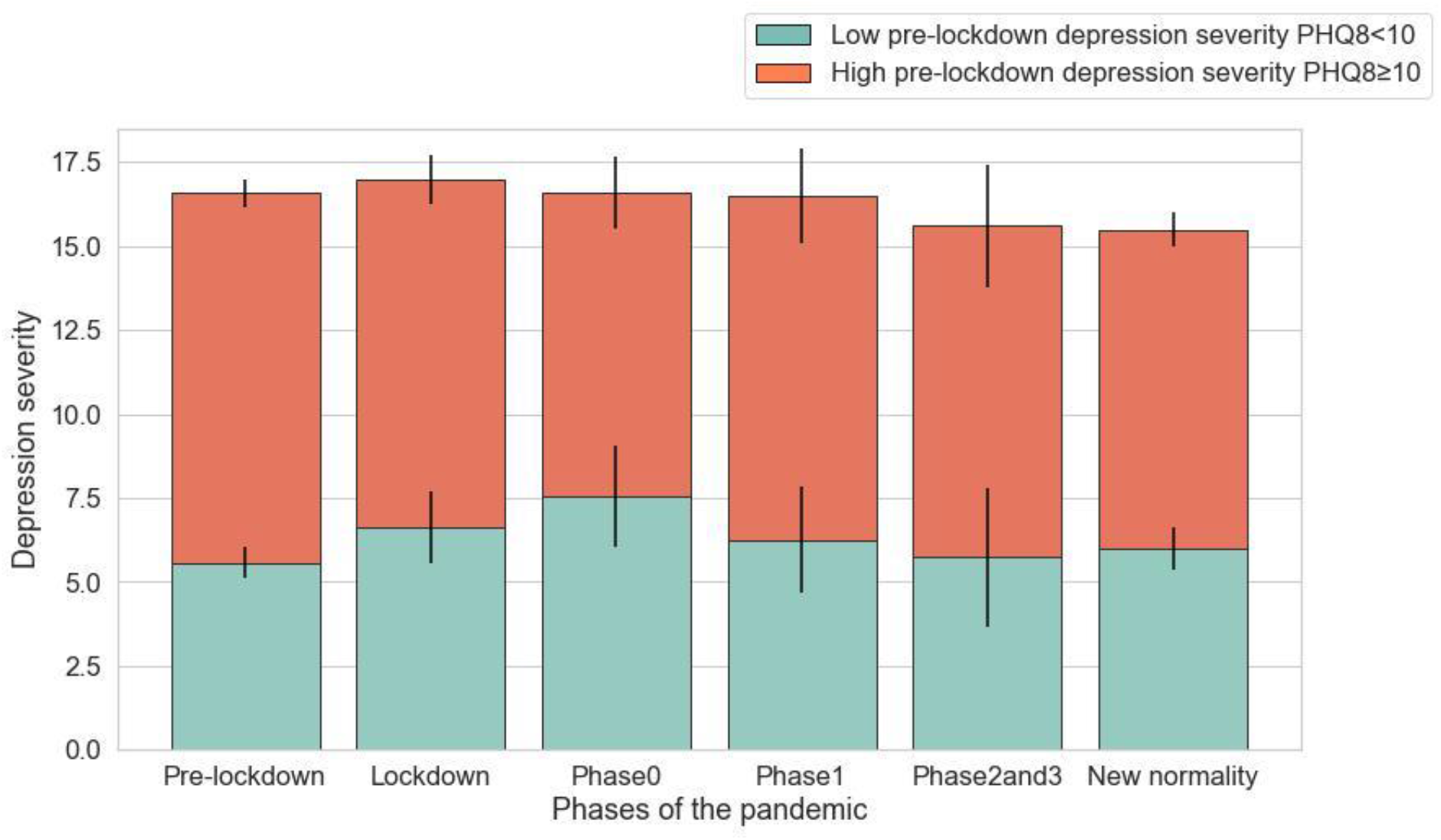
Mean levels and confidence interval of depression (PHQ-8) by pre-lockdown depression severity during each phase. Based on a descriptive analysis, we divided the participants depending on the pre-lockdown depression severity.

**Figure 4.**
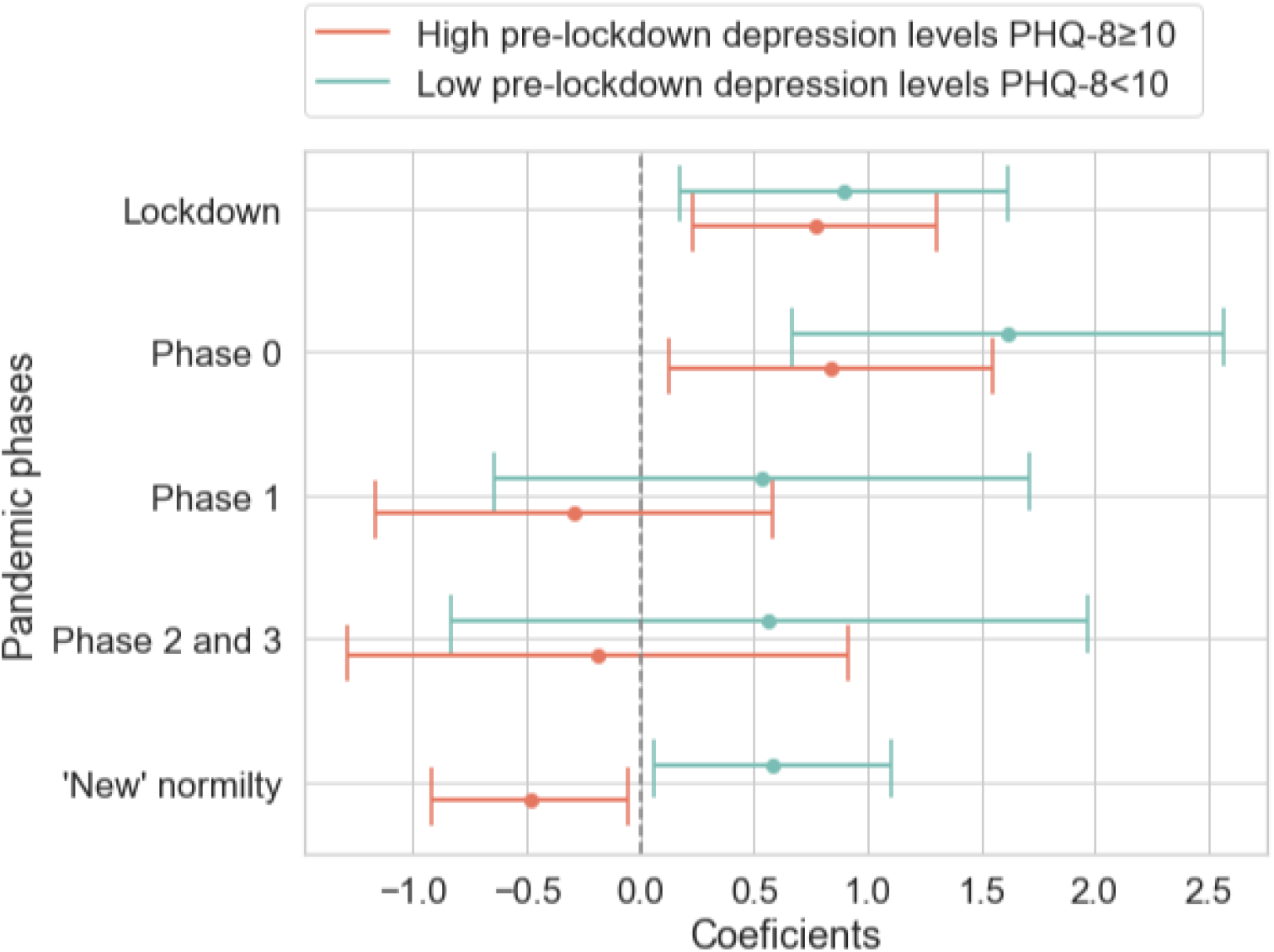
Coefficients of stratification model of pre-lockdown depression. depictured the coefficient of the model (Phase vs pre-lockdown depression levels). Adjusted for the covariates: gender, age, income and number of people living with, and comorbidity with other medical conditions.

Those with high pre-lockdown severity displayed a significant increase in depression severity during the lockdown (β = 0.768, CI 95% [0.227-1.309], *p*≤0.001) and phase 0 (β = 0.838, CI 95% [0.127-1.550], *p=*0.021), as compared to the pre-lockdown severity. Moreover, we found a significant decrease in depression levels during the “new-normality” (β = -0.486, CI 95% [-0.917 - -0.055], *p*=0.027).

As regards the others with low pre-lockdown depression severity, participants also displayed a significant increase in their depression severity during the lockdown (β = 0.894, CI 95% [0.174-1.614], *p*=0.015) and phase 0 (β = 1.615, CI 95% [0.664-2.565], *p*≤0.001), as compared with the pre-lockdown, consistent with the previous model. Instead, we found a significant increase in depression levels during the “new-normality” (β = 0.581, CI 95% [0.056 – 1.105], *p*=0.03). The coefficients of the stratified model adjusted for the covariates are represented in Figure 4.

## DISCUSSION

We observed that the participants with a history of MDD were mainly affected during the lockdown and the early phase of post-lockdown, when their depressive symptoms increased as compared to pre-lockdown levels. These findings should not be surprising, since the most stringent measures were imposed during these phases.

In previous literature, some works (González-Blanco et al., 2020; Solé et al., 2021), showed that patients with mental illness were more depressed during the post-lockdown phase than healthy controls as expected. Another study (García-Álvarez et al., 2020) observed a predominantly depressive response to lockdown in all the participants with a current or a past mental disorder, but also in healthy controls. What is clear is that none of these studies, all of whom conducted in samples of Spanish people, dealt with the specific impact of the pandemic on the depressive symptoms of patients suffering from MDD, unlike two works (Hamm et al., 2020; Quittkat et al., 2020) outside Spain which did. *Quittkat et al (2020)* showed that in depressed patients from Germany depressive symptoms increased during the lockdown, as compared with the pre-lockdown (November 2019). During this lockdown, strict restrictions were imposed, corresponding, in essence, to those existing in Catalonia during the lockdown, although the lockdown in Spain was more stringent than in Germany. This is clearly shown by the data given by the Oxford COVID-19 government response tracker (*OxCGRT*, n.d.): an index called *stringency level* was introduced to describe numerically the severity of restrictions, based on values given to many parameters measuring it, such as the closure of the school, the limitations imposed to internal movements etc. For what concerns April 2020, Spain had a *stringency level* of 85.19, whereas Germany had 76.85. In contrast, the study by Hamm et al. (Hamm et al., 2020) in four American metropolitan areas, provided evidence contrary to our results, as they did not find significant changes in depression in their cohort with MDD, between pre-lockdown and lockdown. However, this divergence of findings can be explained by a far lower stringency level of restrictions (*stringency level* (*OxCGRT*, n.d.) of 72.69 in April 2020). Moreover, analyses were conducted on a sample older than ours (the mean age was 69), thus we might also interpret this apparent lesser impact of social restrictions on the elderly due to a weaker habit of going out than younger people.

Previous studies of individuals with MDD have shown contradictory results in terms of the impact of lockdown and subsequent public health restrictions. As in our sample, a German (Quittkat et al., 2020) study showed an increase in symptoms, another from USA (Hamm et al., 2020) did not. These differences may be attributable to different study designs (single as opposed to repeat measures of depression), different samples or differences in the lockdown regime.

Furthermore, other studies (Leightley et al., 2021) were “lockdown-centric”, since the period in study was divided into a pre-lockdown phase, lockdown, and a post-lockdown phase including indiscriminately the whole period of time following the lockdown. Instead, the present manuscript considered the post-lockdown dividing it into different periods based on the gradual ease of restrictions.

Second, the model suggests that in the “new” normality period those subjects with high pre-lockdown depression experienced a decrease in depressive symptoms.

These results come out as partly divergent from those found in previous studies, but these discrepancies might again rely on the above-discussed differences in the division into phases of our common period of interest. Indeed, *Leightley et al* (2021) included in their post-lockdown phase also the period that in the current study was studied as the phase 1 of the post-lockdown, when some social restrictions were still in place. Thus, as we based our analyses on different divisions of time, slight differences in findings are understandable. However, many studies (Fleischmann et al., 2021; García-Álvarez et al., 2020; González-Blanco et al., 2020; Hamm et al., 2020; Pan et al., 2021; Quittkat et al., 2020; Robinson et al., 2022; Solé et al., 2021) dealt with the fluctuation of depression during the pandemic in depressed patients, but definitely did not considering the influence of different levels of pre-lockdown depression or anxiety on the subsequent variation of depressive symptoms due to the COVID-19 first wave. Therefore, to the best of our knowledge, our study is the first one that describes in-depth the influence of the pre-pandemic levels of depression in the subsequent oscillations in the depressive symptoms of patients with a history of MDD during the different phases of the pandemic. As explained above, the core of our findings suggests not only that the impact of restrictions in terms of depression worsening was significant during the lockdown and the first phase of post-lockdown, but also that the pre-lockdown levels of depressive symptoms exerted a possible influence on the course of depression during the last phase of the post-lockdown. Indeed, during “new-normality” a significant increase of depression was found in those with lower pre-existing levels of depression as compared to the pre-lockdown, whereas in those with higher levels our results displayed a decrease in depression. Furthermore, our sample only consisted of Catalan people living under the same restrictions at the same time, which makes our study population unprecedently homogeneous, also from a socio-cultural perspective.

These results provide unprecedented points of reflection, soliciting the proposal of possible explanations. There is no doubt that the COVID-19 pandemic produced a contraction of the habitual interactions of the individual with the stimuli produced by the external world. Thus, we might suppose that the psychological impact of such everyday life upheaval must have been more critical on those patients whose pre-lockdown depressive and anxiety symptoms were not severe to such an extent that their psychological wellbeing could be influenced by relevant changes in the external world. In other words, high levels of depression before the pandemic might have made individuals less responsive to the stress (Kontaxis et al., 2021), and prevented them from suffering from a persistent increase in depressive symptoms during the periods of social restrictions (Osório et al., 2017). However, given the scarcity of past references on the theme, these hypotheses need to be further proved by future analyses on the possible role of pre-pandemic levels of depression and anxiety as predictors for the impact of severe social restrictions on the course of depressive symptoms in patients affected by MDD.

### Implications and future directions

First of all, the study of the effects of restrictions on depressive symptoms in a clinical population of people suffering from MDD has to be regarded as providing a relevant clinical implication. The results of the current work about the specific psychological impact of the different measures imposed a long time, could inform authorities in planning a future policy of restrictions in the case of new pandemics, taking into account more accurately and from several points of view the benefits and disadvantages of applying social restrictions.

For example, analyses on our sample suggest that during the lockdown and the early phase of the post-lockdown, when only some outdoor sport activities were allowed, the levels of depression severity remained high, whereas a drop was registered during phase 1, when some restrictions of social activities were slightly eased. Thus, it is hoped that, in case of future pandemics, this specific vulnerability to restrictions that we described in depressed patients will induce authorities to, for example, exempt them from the strictest forms of lockdown, compatibly with the epidemiological context.

### Limitations and Strengths

Some limitations have to be considered. The assessment of severity was self-reported and may have lower reliability than heteroadministered (administered by a clinician on behalf of a patient) questionnaires or tests. Furthermore, some subjects missed data referring to certain phases.

Strengths Of the study include a homogeneous sample in terms of geographical origin and we were able to analyze lockdown measures in detail. Since we considered the differences between different areas within Catalonia, each group of patients corresponding to a specific phase contained only participants who were imposed under the same restrictions at the same time. Moreover, thanks to the length of our period of assessment, for each participant several observations have been available, referring to the pre-lockdown, lockdown, and post-lockdown phases. Thus, the present research went beyond the “lockdown-centric” perspective of the previous literature and has to be considered the first one assuming a “pandemic-centric” point of view, which not only considered the acute impact of the most severe restrictions but also aimed at detecting the subtle variations in depression during the post-lockdown phases.

Furthermore, due to the employment of RMT and the prospectively-planned, longitudinal design of our study, recall bias was completely prevented. Also, our database allowed us to distinctly study the role of pre-lockdown levels of depression and anxiety as predictors for variations in depressive symptoms during lockdown and post-lockdown.

### Conclusion

Future studies will have the possibility to refer to the present work as a significant milestone of this new “pandemic-centric” perspective, based on which not only an impairment in depression severity during the first phases of pandemic was found, but also a more severe impact of the pandemic was described on those patients suffering from milder forms of MDD. Thus, our results emphasize the strong need for attentive care that these specific populations run under social restrictions, hopefully contributing to the process of turning the spotlight on the psychological impact of the pandemics.

## Data Availability

All data produced in the present study are available upon reasonable request to the authors

## CRediT authorship contribution statement

According to the Contributor Roles Taxonomy (CRediT; https://credit.niso.org/), each author’s role in the current manuscript are summarised below:

*Conceptualization*: Lavalle R., Condominas E., Siddi S. Haro JM.

*Data curation and formal analysis*:, Condominas E.

*Funding Acquisition:* Hotopf M., Narayan V.

*Investigation:* F Matcham, F Lamers, S Siddi, P Annas, KM White, C Oetzmann, BWJH Penninx, JM Haro, M Hotopf

*Methodology:* Lavalle R., Condominas E., Siddi S., Garcia E., Giné-Vázquez I., Laporta E., Bailon R, Kontaxis S

*Project Administration and resources:* F Matcham, F Lamers, S Siddi, BWJH Penninx, JM Haro, M Hotopf

*Software:* Condominas E, Giné-Vázquez I., Laporta E.

*Supervision*; F Matcham, F Lamers, S Siddi, P Annas, JM Haro, S Simblett, M Hotopf Validation: Siddi S., Haro JM

Visualisation: Lavalle R., Condominas E., Siddi S.

Writing – original draft: Lavalle R., Condominas E., Siddi S.

Writing – review and editing: all authors.

## Acknowledgment

The RADAR-CNS project has received funding from the Innovative Medicines Initiative 2 Joint Undertaking under grant agreement No 115902. This Joint Undertaking receives support from the European Union’s Horizon 2020 research and innovation programmer and EFPIA, www.imi.europa.eu. This communication reflects the views of the RADAR-CNS consortium and neither IMI nor the European Union and EFPIA are liable for any use that may be made of the information contained herein. The funding body have not been involved in the design of the study, the collection or analysis of data, or the interpretation of data.

Participants in the CIBER site came from following four clinical communities in Spain: Parc Sanitari Sant Joan de Déu Network services, Institut Català de la Salut, Institut Pere Mata, and Hospital Clínico San Carlos.

This paper represents independent research part funded by the National Institute for Health Research (NIHR) Maudsley Biomedical Research Centre at South London and Maudsley NHS Foundation Trust and King’s College London. The views expressed are those of the author(s) and not necessarily those of the NHS, the NIHR or the Department of Health and Social Care. We thank all the members of the RADAR-CNS patient advisory board for their precious contribution to the procedures of device selection, and their helpful advice throughout the study protocol design. This research was reviewed by a team with experience of mental health problems and their careers who have been specially trained to advise on research proposals and documentation through the Feasibility and Acceptability Support Team for Researchers (FAST-R): a free, confidential service in England provided by the National Institute for Health Research Maudsley Biomedical Research Centre via King’s College London and South London and Maudsley NHS Foundation Trust. The views expressed are those of the authors and not necessarily those of the NHS, the NIHR, or the Department of Health and Social Care. NIHR Senior Investigator Awards have supported Matthew Hotopf.

## Abbreviations

aRMT: active remote measurement technology
GAD-7: item Generalized anxiety disorder-7 item
IQR: Interquartile ranges
MDD: Major depressive disorder
ML: maximum likelihood
pRMT: passive remote measurement technology
PHQ-8: Patient health questionnaire 8-item
RADAR-CNS: Remote assessment of disease and relapse - central nervous system
REDCap: Research electronic data capture
RADAR-MDD: Remote Assessment of Disease and Relapse-Major Depressive Disorder

