## Supplemental document 1 for "The impact of COVID-19 lockdown on a cohort of adults with recurrent major depressive disorder from Catalonia: a decentralized longitudinal study using remote measurement technology"

***Document 1. Description of the lockdown phases.***

Different phases of restrictions in Catalonia, based on the restrictions of the three studied districts:

- Pre-lockdown (from November 1, 2019, to March 11, 2020): for this period we chose November 1 as the starting date until March 11. Indeed, on this last day, the Government of Catalonia introduced social distancing to fight the spread of COVID-19 (RESOLUCIÓ SLT/704/2020, d'11 de Març, 2020)

- Lockdown (from March 11, 2020, to April 27, 2020): on March 14, 2020 (Real Decreto 463/2020, de 14 de Marzo, BOE n.°3692, 2020) was declared the State of Alarm, and the Spanish population was subjected to a severe lockdown from the following day. Mobility was limited to essential activities and educational institutions were closed (García-Esquinas et al., 2021). Then, the Catalan authorities introduced supplementary measures, on March 12, 13, 18, 23 (RESOLUCIÓ SLT/719/2020, de 12 de Març, 2020; RESOLUCIÓ SLT/720/2020, de 13 de Març, 2020; RESOLUCIÓ SLT/746/2020, de 18 de Març, 2020; RESOLUCIÓ SLT/761/2020, de 23 de Març, 2020), which produced the suspension of any in-person educational activity, the closure of libraries and museums, a temporary reorganization of the health system, etc.

Later restrictions were lifted gradually through four phases of the post-lockdown:

- Phase 0 (from April 27 2020 to May 10 or 17 or 24, 2020 depending on the district): restrictions began to be lifted on May 4 in all of Spain (Disposición 4791, BOE n.°123, 2020; Disposición 7351, BOE n.°107, 2021), when non-essential businesses were opened by appointment and citizens were allowed to do outdoor physical activities (1h a day maximum) by time slot based on age. In this study, we considered April 27 as the beginning of phase 0, since the relaxation of confinement and the permission for children to go for a walk had already been instituted seven days earlier than May 4

- Phase 1 (from May 11 or 18 or 25 2020 to May 24 or June 7, depending on the district): (Ajuntament de Barcelona, 2020b; Disposición 4911, BOE n.°130, 2020; Orden SND/399/2020, de 9 de Mayo, BOE n.°4911, 2020; la CIUTAT Diari digital de proximitat, 2020) measures were gradually relaxed: meetings with a maximum of 10 people were allowed; the bars and restaurants opened (only outdoor space), as well as some spaces of culture, museums, and gyms; transfers to a second residence were permitted.

- Phases 2 and 3 (from May 25 or June 8 2020 to June 18 depending on the district): (Ajuntament de Barcelona, 2020a; ELNACIONAL.CAT., 2020; Jefatura de Estado, 2020; SER, 2020a)time slots were finished and bars and restaurants' openings were extended even to the indoor areas, with limited capacity. Shopping centers opened, public transport restarted working at 100%, and the percentage of capacity in cinemas, theaters, and museums increased. Phase 3 slightly differed from the previous one: the main discrepancies consisted in a higher capacity allowed for indoor activities and in the fact that ceremonies could again take place. Phases 2 and 3 were considered together, as the latter was very short, for example, 2 days in total in Barcelona (betevé, 2020; SER, 2020b), and differences among them were minimal.

- New normality (from June 19 to October 16): on June 18 the Catalan authorities signed the *resoluciò INT/1433/2020* (RESOLUCIÓ INT/1433/2020, de 18 de Juny, 2020): finally, the Spanish population could experience a new normality. The only resisting restrictions concerned social distancing, the use of a mask, avoidance of crowds of people, and maintenance of some capacity limitations. October 16, 2020, was chosen as the end of the phase when there was a resurgence of COVID-19 and the government set in place new restrictions (Disposición 11590, BOE n.°260, 2020).
