## Supplemental Table 1 for "The impact of COVID-19 lockdown on a cohort of adults with recurrent major depressive disorder from Catalonia: a decentralized longitudinal study using remote measurement technology"

***Table S1. Dates of each phase of pandemic in Catalonia, based on the three studies distructs*.**

| Phase of pandemic | Distructs | Starting day | Ending day |
| --- | --- | --- | --- |
| *Pre- lockdown* | BAR, TAR and GAR | 01/11/2019 | 10/03/2020 |
| *Lockdown* | BAR, TAR and GAR | 11/03/2020 | 26/04/2020 |
| *Phase 0* | BAR | 27/04/2020 | 24/05/2020 |
| | TAR | 27/04/2020 | 10/05/2020 |
| | GAR | 27/04/2020 | 17/05/2020 |
| *Phase1* | BAR | 25/05/2020 | 07/06/2020 |
| | TAR | 11/05/2020 | 24/05/2020 |
| | GAR | 18/05/2020 | 07/06/2020 |
| *Phase 2 and 3* | | | |
| | BAR | 08/06/2020 | 18/06/2020 |
| | TAR | 25/05/2020 | 18/06/2020 |
| | GAR | 08/06/2020 | 18/06/2020 |
| *New normality* | BAR, TAR and GAR | 19/06/2020 | 16/10/2020 |

***Note**: BAR: Barcelona, TAR: Tarregona, GAR: Garraf.*
