## Supplemental Table2 for "The impact of COVID-19 lockdown on a cohort of adults with recurrent major depressive disorder from Catalonia: a decentralized longitudinal study using remote measurement technology"

***Table S2.*** ***Number of participants, observations and average PHQ-8 values in each phase***

| **Phases** | **Observations** | **N** | **PHQ8** | | |
|---|---|---|---|---|---|
| | | | **Mean** | **CI (95%)** | **Median** |
| ***Pre-lockdown*** | 659 | 121 | 12.788 | [12.28 to 13.29] | 13 |
| ***Lockdown*** | 215 | 88 | 13.763 | [12.88 to 14.64] | 15 |
| ***Phase 0*** | 109 | 72 | 13.688 | [12.51 to 14.86] | 14 |
| ***Phase 1*** | 66 | 63 | 13.227 | [11.64 to 14.81] | 14 |
| ***Phases 2-3*** | 43 | 40 | 12.163 | [10.18 to 14.15] | 12 |
| ***New normality*** | 503 | 84 | 12.048 | [11.49 to 12.61] | 12 |

***Note:*** *The observations are the PHQ-8 assessments of each phase. N corresponds to the number of people of those 121 who have registration in the corresponding phase.*
